# Prediction of Colorectal Cancer Risk Based on Profiling with Common Genetic Variants

**DOI:** 10.1101/19010116

**Authors:** Xue Li, Maria Timofeeva, Athina Spiliopoulou, Paul McKeigue, Yazhou He, Xiaomeng Zhang, Victoria Svinti, Harry Campbell, Richard S Houlston, Ian PM Tomlinson, Susan M Farrington, Malcolm G Dunlop, Evropi Theodoratou

## Abstract

**Background:** Stratifying the risk of colorectal cancer (CRC) based on polygenic risk scores (PRSs) within populations has the potential to optimize screening and develop targeted prevention strategies.

**Methods:** A meta-analysis of eleven genome-wide association studies (GWAS), comprising 16 871 cases and 26 328 controls, was performed to capture CRC susceptibility variants. Genetic models with several candidate PRSs were generated from Scottish CRC case–control studies (6478 cases and 11 043 controls) for prediction of overall and site-specific CRC. Model performance was validated in UK Biobank (4800 cases and 20 287 controls). The 10-year absolute risk of CRC was estimated by modelling PRS with age and sex using the CRC incidence and mortality rates in the UK population.

**Findings:** A weighted PRS including 116 CRC SNPs (wPRS_116_) showed the strongest performance. Deconstructing the PRS into multiple genetic risk regional scores or inclusion of additional SNPs that did not reach genome-wide significance did not provide any further improvement on predictive performance. The odds ratio (OR) for CRC risk per SD of wPRS_116_ in Scottish dataset was 1·46 (95%CI: 1·41-1·50, c-statistics: 0·603). Consistent estimates were observed in UK Biobank (OR=1·49, 95%CI: 1·44-1·54, c-statistics: 0·610) and showed no substantial heterogeneity among tumor sites. Compared to the middle quintile, those in the highest 1% of PRSs had 3·25-fold higher risk and those in the lowest 1% had 0·32-fold lower risk of developing CRC. Modelling PRS with age and sex in the general UK population allows the identification of a high-risk group with 10-year absolute risk ≥5%.

**Interpretation:** By optimizing wPRS_116_, we show that genetic factors increase predictive performance but this increment is equivalent to the extraction of only one-tenth of the genetic susceptibility. When employing genetic risk profiling in population settings it provides a degree of risk discrimination that could, in principle, be integrated into population-based screening programs.

## Research in context

### Evidence before this study

The recent progress in genome-wide association studies (GWASs) has discovered an increasing number of CRC-associated risk variants. Stratifying population risk of colorectal cancer (CRC) based on polygenic risk scores (PRSs) could improve screening and prevention strategies.

### Added value of this study

Our study found that a PRS including 116 CRC susceptibility SNPs (wPRS_116_) was the optimal score, while deconstructing genetic risk into multiple regional scores or inclusion of additional SNPs above genome-wide significance threshold showed no further improvement on prediction performance. By optimizing wPRS_116_, we show that genetic factors in combination with age, sex and family history increase predictive performance but this increment is small, equivalent to only about one-tenth of the genetic information that could be extracted by the optimal polygenic score. In population settings, employing genetic risk profiling can achieve a degree of risk discrimination that is useful to inform the optimal design of population-based screening programs.

### Implications of all the available evidence

It is expected that those identified to be at higher CRC risk by their genetic profile of wPRS_116_ might be more likely to participate in screening, thereby further increasing the screening uptake and detection rates. Implementation of PRS-based risk profiling in conjunction with quantitative fecal immunochemical test (qFIT) based screening would achieve a fine-tuned sensitivity/specificity of the qFIT test. The application of a high cut-off qFIT threshold in those at low genetic risk could avoid invasive tests and minimize adverse effects and cost.

## INTRODUCTION

Colorectal cancer (CRC) is one of the most common cancers, with 1·8 million new cases and almost 0·8 million deaths globally in 2018.^1^ Substantial evidence showed that screening can reduce the CRC mortality by allowing early detection and removal of precancerous lesions.^2^ Policy makers and clinicians rely on risk classification to determine which individuals to screen. To date, these classification schemes are predominantly based on age and/or a simple classification of family history. Stratifying the average risk population into risk categories offers the potential of tailoring surveillance intensity, through which individuals at high risk could benefit from earlier or more frequent screening, whereas others at low risk could delay the onset or reduce the frequency of screening.

Comprehensive information on genetic susceptibility could contribute importantly to CRC risk stratification, given that the heritability of CRC has been estimated to be around 16%-35%^3^ and the sibling recurrence risk ratio is about 2·0.^4^ We previously assessed the utility of CRC genetic risk profiling with a panel of 10 common genetic variants associated with CRC susceptibility.^5^ Although discrimination ability was low (c-statistic of 0·56), we showed that genotype data provides additional information to that from family history alone.^5^ Others have also showed that personalized screening using polygenic risk scores (PRSs) have the potential to optimize population screening for CRC and could identify subgroups most likely to benefit from targeted CRC prevention strategies.^6^ Incorporating more complete genetic information is expected to improve the risk stratification and the combined effect of multiple risk loci has the potential to achieve a degree of risk discrimination that is useful for population-based prevention and screening programs.

The recent progress in genome-wide association studies (GWASs) has discovered an increasing number of CRC-associated risk variants. These findings provide further insight into CRC susceptibility and enhance the prospects of applying genetic risk scores to both personalized and population-based screening and prevention. In this study, we aim to develop CRC prediction models and to assess model performance in both individual and population settings. We developed models by incorporating the genetic information of CRC and several markers that comprise potential CRC risk factors or complex traits co-occurring with CRC. To gauge the broader future potential of genetic risk modelling, we assessed the utility of genetic risk scores in categorizing risk subgroups within the general population by projecting the risk models to the UK population.

## METHODS

### Studies

We made use of 11 previously published GWASs (i.e., CCRR1, CCFR2, COIN, CORSA, Croatia, DACHS, FIN, NSCCG-OncoArray, SCOT, UK1 and VQ58) to generate a list of genetic variants associated with CRC risk.^7-17^ A series of Scottish CRC case–control studies were used to test the predictive performance of polygenic risk scores (PRSs). The developed PRSs were further evaluated in an independent test dataset from UK Biobank. Standard quality control (QC) measures were applied to each of the datasets. After the QC process, a total of 16 871 cases and 26 328 controls were finally included for the derivation of genetic susceptibility SNPs, 6478 cases and 11 043 controls from the Scottish dataset were included for the validation of PRSs, and 4800 cases and 20 287 controls from the UK Biobank were included to further test the optimized PRSs. Details of these studies are described in appendix p 1 Supplementary Methods and appendix p 2 Table S1.

### Polygenic risk scores

#### Genome-wide polygenic score

We performed a meta-GWAS of 11 studies to obtain a list of genome-wide significant SNPs (p<5×10^−8^) and their per-allele odds ratios (ORs) and standard errors for CRC risk. For completeness, we also included the genetic risk variants reported in early published CRC GWASs (appendix p 3 Table S2). A weighted genome-wide PRS (*w*PRS) was computed using both previously known susceptibility variants and independent variants identified by the meta-GWAS.

#### Regional genetic scores

By using only the genome-wide significant SNP, some correlated informative signals that are independently associated with CRC may be excluded. Therefore, we constructed regional genetic scores by including SNPs associated with CRC and its risk factors (i.e., vitamin D [VD], C-reactive protein [CRP], body mass index [BMI], waist hip rate [WHR], and inflammatory bowel disease [IBD]). Regional genetic scores were calculated by using the GENOSCORES library (https://pm2.phs.ed.ac.uk/genoscores/). This is similar to the approach used for LDpred,^18^ in which the correction for LD between SNPs was based on pre-multiplying the vector of weights by the generalized inverse of the correlation matrix estimated from 1000G reference panel of European ancestry.

### Model development and evaluation

We constructed prediction models in the Scottish dataset by incorporating genetic CRC risk in the form of either PRSs or regional genetic scores with adjustment for the first 10 genetic principal components (PCs). A sequence of logistic models was fitted for: (i) a weighted PRS of identified CRC GWAS SNPs; (ii) regional genetic scores for CRC; and (iii) regional genetic scores for CRC and other relevant traits. A series of stepwise backward logistic regressions was conducted on regional genetic scores to obtain an optimized set of scores determined by the Akaike information criterion (AIC). The discriminatory accuracy of the developed models was evaluated by the area under the receiver-operating characteristic curve (known as c-statistic) with 10-fold cross-validation. These models were further stratified by anatomic tumor sites (i.e., proximal colon, distal colon and rectum). The PRS models with the best performance in Scottish dataset were evaluated in UK Biobank in terms of discrimination and calibration. Odd ratios (ORs) were then derived per SD increase in PRS for overall, and site-specific, CRC risk. To simplify the interpretation of PRS, we categorized it into percentiles based on its distribution in controls.

### Combined effect of PRS and family history

To evaluate the incremental contribution of combining PRS and family history for prediction, we additionally calculated the expected information for discrimination (expected weight of evidence, denoted as Λ).^19^ Briefly, the expected information for discrimination is the expected log-likelihood ratio in favor of correct assignment as case or control, taken as the average of the value in cases and the value in controls. One advantage of using Λ is that the contributions of independent variables to predictive performance are additive on the scale of Λ. For a logistic regression model, the sampling distribution of Λ is asymptotically Gaussian. In this situation, the c-statistic can be viewed as a mapping of Λ, which takes values from 0 to infinity to the interval from 0·5 to 1.^20^ We then recalibrated the posterior probabilities by fitting a logistic regression model with the outcome as response variable and the logit of the posterior probability as the predictive variable.

### Estimation of absolute risk for developing CRC

The absolute risk of CRC for individuals in each risk category (i.e. each percentile of PRS) was calculated after accounting for competing risks of dying from causes other than CRC by using the formula described previously.^21^ Specifically, we obtained sex- and age-dependent UK CRC incidence and mortality rates for 2016 mid-year from the Office for National Statistics (http://www.ons.gov.uk/). The mortality rates for non-CRC causes were estimated by subtracting the age- and sex-specific CRC mortality rates from the overall mortality rates. Full details of these calculations are provided in appendix p 1 Supplementary Methods.

## RESULTS

### Deriving CRC susceptibility SNPs and creating polygenic risk scores

The meta-analysis of 11 GWASs resulted in the identification of 1593 genetic variants that were associated with CRC at p<5×10^−8^. After adding SNPs reported in other GWASs and after excluding SNPs in LD, a list of 116 SNPs (appendix p 3 Table S2) were retained for the creation of weighted genome-wide polygenic risk score (wPRS_116_).

We additionally created 35 regional genetic scores that included 1593 SNPs with p<5×10^−8^ (Table 1). We also used more liberal p-value thresholds and created 40 genetic scores comprising of 1837 SNPs at p<10^−7^ and 41 genetic scores comprising of 2712 SNPs at p<10^−6^. The genes harbored in these genomic regions were annotated and are presented in appendix p 4 Table S3. We additionally created 17 regional scores for CRP, 5 for VD, 85 for IBD, 69 for BMI and 48 for WHR with p-value threshold setting as 5×10^−8^. More liberal p-value thresholds (p<10^−7^ and p<10^−6^) were also applied for these traits, and the number of regional genetic scores created and SNPs included are present in Table 1 and Table 2.

**Table 1.** Comparison of methods for deriving the genetic scores: Results from Scottish dataset (SOCCS).

| SNPs selection | SNPs included for creation of scores | Scores entering model (n) | CRC overall |  | Proximal |  | Distal |  | Rectal |  |
| --- | --- | --- | --- | --- | --- | --- | --- | --- | --- | --- |
|  |  |  | Scores selected (n) | C-statistics | Scores selected (n) | C-statistics | Scores selected (n) | C-statistics | Scores selected (n) | C-statistics |
| Genome-wide significant SNPs |  |  |  |  |  |  |  |  |  |  |
| CRC GWAS SNPs | 116 | One weighted PRS | 1 | 0.603 | 1 | 0.562 | 1 | 0.587 | 1 | 0.587 |
| Stepwise regression of CRC LD-adjusted regional scores with varying p-value thresholds |  |  |  |  |  |  |  |  |  |  |
| < 5×10<sup>-8</sup> | 1593 | 35 scores | 35 | 0.584 | 16 | 0.583 | 21 | 0.584 | 23 | 0.586 |
| <10<sup>-7</sup> | 1837 | 40 scores | 36 | 0.584 | 19 | 0.585 | 23 | 0.587 | 25 | 0.587 |
| <10<sup>-6</sup> | 2712 | 41 scores | 36 | 0.586 | 19 | 0.589 | 25 | 0.590 | 26 | 0.591 |

\* C-statistics were estimated from the 10-fold cross-validation

**Table 2.** Stepwise regression of LD-adjusted regional scores for CRC and related traits with varying p-value thresholds (SOCCS).

| SNPs selection | SNPs included for creation of scores | Scores entering model (n) | Scores selected by the model (n) | C-statistics* |
| --- | --- | --- | --- | --- |
| $< 5 \times 10^{-8}$ | 1593 CRC SNPs + 870 CRP SNPs + 521 VD SNPs + 3217 UC SNPs + 4814 CD SNPs + 1501 BMI SNPs + 708 WHR SNPs | 35 CRC scores + 17 CRP scores + 5 VD scores + 35 CD scores + 50 UC scores + 69 BMI scores + 39 WHR scores | 31 CRC scores + 7 CRP scores + 2 VD scores + 25 IBD scores + 18 BMI scores + 7 WHR scores | Overall: 0.595<br>Proximal: 0.594<br>Distal: 0.585<br>Rectal: 0.583 |
| $< 10^{-7}$ | 1837 CRC SNPs + 911 CRP SNPs + 540 VD SNPs + 3464 UC SNPs + 5168 CD SNPs + 1622 BMI SNPs + 811 WHR SNPs | 40 CRC scores + 17 CRP scores + 6 VD scores + 39 CD scores + 50 UC scores + 74 BMI scores + 48 WHR scores | 35 CRC scores + 6 CRP scores + 2 VD scores + 26 IBD scores + 20 BMI scores + 11 WHR scores | Overall: 0.596<br>Proximal: 0.597<br>Distal: 0.589<br>Rectal: 0.585 |
| $< 10^{-6}$ | 2,712 CRC SNPs + 1,152 CRP SNPs + 656 VD SNPs + 4899 UC SNPs + 6690 CD SNPs + 2359 BMI SNPs + 1178 WHR SNPs | 41 CRC scores + 18 CRP scores + 7 VD scores + 40 CD scores + 51 UC scores + 76 BMI scores + 49 WHR scores | 35 CRC scores + 6 CRP scores + 1 VD scores + 31 IBD scores + 16 BMI scores + 9 WHR scores | Overall: 0.598<br>Proximal: 0.593<br>Distal: 0.595<br>Rectal: 0.586 |

\* C-statistics were estimated from the 10-fold cross-validation

### Optimizing the PRSs for overall and site-specific CRC risk

We set out to validate these derived scores and to choose the best score for further analysis by examining their discrimination in a Scottish dataset (appendix p 5 Table S4). Specially, the combined effect of 116 CRC SNPs in the form of wPRS_116_ was statistically significantly associated with CRC risk (OR=1·46, 95% CI: 1·41-1·50, p=1·71×10^−116^, 1 SD increase of wPRS_116_) (Table 3). The ORs per 1 SD increase of the wPRS_116_ were 1·26 (95% CI: 1·19-1·33, p=1·47×10^−17^) for the proximal, 1·36 (95% CI: 1·30-1·45, p=3·46×10^−29^) for the distal and 1·37 (95% CI: 1·30-1·44, p=6·37×10^−37^) for the rectum. The c-statistic for the predictive ability of wPRS_116_ was 0·603, showing moderate discrimination. Calibration of the predictive model of wPRS_116_ is shown in appendix p 6 Figure S1. When stratifying the CRC status by tumor sites, the predictive ability of wPRS_116_ for cancer at proximal colon (c-statistic=0·562), distal colon (c-statistic=0·587) and rectum (c-statistic=0·587) had less accuracy than that for overall CRC risk.

**Table 3:** The association between wPRS_116_ and CRC risk in SOCCS and UKBB.

| Tumor sites | Validation dataset (SOCCS) |  |  | Test dataset (UKBB) |  |  |
| --- | --- | --- | --- | --- | --- | --- |
|  | OR (95% CI) | P | C-statistics* | OR (95% CI) | P | C-statistics * |
| CRC overall | 1.46 (1.41-1.50) | 1.71×10<sup>-116</sup> | 0.603 | 1.49 (1.44-1.54) | 6.67×10<sup>-128</sup> | 0.610 |
| Proximal | 1.26 (1.19-1.33) | 1.47×10<sup>-17</sup> | 0.562 | 1.51 (1.43-1.61) | 6.00×10<sup>-42</sup> | 0.614 |
| Distal | 1.36 (1.30-1.45) | 3.46×10<sup>-29</sup> | 0.587 | 1.48 (1.41-1.55) | 1.25×10<sup>-54</sup> | 0.607 |
| Rectal | 1.37 (1.30-1.44) | 6.37×10<sup>-37</sup> | 0.587 | 1.47 (1.39-1.55) | 6.73×10<sup>-41</sup> | 0.606 |

\* C-statistics were estimated from the 10-fold cross-validation

To minimize the cost of adding noise from the inclusion of multiple regional scores that were not truly associated with CRC risk, we performed stepwise backward regression. The best set of regional scores included 31 CRC scores, 7 CRP scores, 2 VD scores, 25 IBD scores, 18 BMI scores and 7 WHR scores. This yielded a c-statistics of 0·595 (Table 2). When stratifying CRC risk by tumor sites and using them as different endpoints for stepwise regression, the best set of regional scores yielded a c-statistic of 0·594 for proximal cancer, 0·585 for distal cancer and 0·583 for rectal cancer. Comparing to the wPRS_116_, the regional scores showed no further improvement on the prediction of CRC.

### Testing the polygenic risk scores in UK Biobank

Having derived the model of polygenic scores with the best discrimination, we next explored their predictive power in an independent dataset (appendix p 5 Table S4). The wPRS_116_ showed moderate discrimination ability and good calibration in UK Biobank: the c-statistic was 0·610 and the calibration plot is shown in appendix p 7 Figure S2. When assessing the predictive ability of wPRS_116_ after tumor site stratification, it yielded a c-statistic of 0·614 for proximal, 0·605 for distal and 0·606 for rectal. The best set of regional scores developed for site-specific CRC in Sottish dataset did not outperform the wPRS_116_ in UK Biobank dataset therefore the remaining analysis concentrated on the wPRS_116_ for simplicity.

The ORs per 1 SD increase of the wPRS_116_ were 1·49 (95% CI: 1·44-1·54, p=6·67×10^−128^) for overall CRC, 1·51 (95% CI: 1·43-1·61, p=6×10^−42^) for proximal, 1·48 (95% CI: 1·41-1·55, p=1·25×10^−54^) for distal and 1·47 (95%CI: 1·39-1·55, p=6·73×10^−41^) for rectal (Table 4). For individuals in the lowest 1% of wPRS_116_, the OR compared with the middle quintile (40%-60%) was 0·32 (95%CI: 0·19-0·54, p=8·51×10^−6^). By contrast, for individuals in the highest 1% of the PRS distribution, the corresponding estimated OR was 3·25 (95%CI: 2·50-4·22, p=1·52×10^−17^). When considering CRC risk separately for proximal colon, distal colon and rectum, the corresponding ORs showed no substantial heterogeneity.

**Table 4.** Association between the percentiles of wPRS_116_ and CRC risk in different sites: Odds ratios and 95% Confidence Intervals (UKBB).

| Allele count percentile | Overall CRC |  |  |  | Proximal |  |  | Distal |  |  | Rectal |  |  | P heterogeneity |
| --- | --- | --- | --- | --- | --- | --- | --- | --- | --- | --- | --- | --- | --- | --- |
|  | N control | N case | OR (95%CI) | P | N case | OR (95%CI) | P | N case | OR (95%CI) | P | N case | OR (95%CI) | P |  |
| <1% | 234 | 16 | 0.32 (0.19 - 0.54) | 8.51×10<sup>-6</sup> | 4 | 0.30 (0.11 - 0.81) | 0.018 | 6 | 0.34 (0.15 - 0.78) | 0.011 | 4 | 0.28 (0.10 - 0.77) | 0.012 | 0.956 |
| 1-5% | 912 | 92 | 0.48 (0.38 - 0.60) | 1.04×10<sup>-10</sup> | 20 | 0.38 (0.24 - 0.61) | 3.56×10<sup>-5</sup> | 33 | 0.49 (0.34 - 0.70) | 1.05×10<sup>-4</sup> | 33 | 0.60 (0.41 - 0.87) | 0.008 | 0.326 |
| 5-10% | 1120 | 135 | 0.57 (0.47 - 0.69) | 1.17×10<sup>-8</sup> | 37 | 0.58 (0.41 - 0.82) | 0.003 | 45 | 0.54 (0.39 - 0.74) | 1.56×10<sup>-4</sup> | 39 | 0.58 (0.41 - 0.81) | 0.002 | 0.941 |
| 10-20% | 2163 | 346 | 0.76 (0.66 - 0.87) | 6.46×10<sup>-5</sup> | 77 | 0.62 (0.48 - 0.81) | 4.43×10<sup>-4</sup> | 137 | 0.85 (0.69 - 1.05) | 0.136 | 99 | 0.76 (0.60 - 0.96) | 0.026 | 0.182 |
| 20-40% | 4206 | 810 | 0.91 (0.82 - 1.01) | 0.093 | 174 | 0.72 (0.59 - 0.88) | 0.002 | 315 | 1.00 (0.85 - 1.18) | 0.993 | 219 | 0.86 (0.72 - 1.04) | 0.132 | 0.044 |
| 40-60% | 4143 | 874 | 1.00 | -- | 237 | 1.00 | -- | 309 | 1.00 | -- | 250 | 1.00 | -- | -- |
| 60-80% | 3882 | 1138 | 1.39 (1.26 - 1.53) | 6.06×10<sup>-11</sup> | 261 | 1.18 (0.98 - 1.41) | 0.089 | 421 | 1.45 (1.25 - 1.70) | 1.82×10<sup>-6</sup> | 318 | 1.36 (1.14 - 1.61) | 5.32×10<sup>-4</sup> | 0.233 |
| 80-90% | 1918 | 589 | 1.46 (1.29 - 1.64) | 4.28×10<sup>-10</sup> | 152 | 1.39 (1.12 - 1.71) | 0.003 | 216 | 1.51 (1.26 - 1.81) | 1.02×10<sup>-5</sup> | 164 | 1.42 (1.16 - 1.74) | 9.53×10<sup>-4</sup> | 0.825 |
| 90-95% | 893 | 362 | 1.92 (1.67 - 2.22) | 4.23×10<sup>-18</sup> | 97 | 1.90 (1.48 - 2.43) | 3.60×10<sup>-7</sup> | 136 | 2.04 (1.65 - 2.53) | 4.71×10<sup>-11</sup> | 94 | 1.74 (1.36 - 2.24) | 1.21×10<sup>-5</sup> | 0.637 |
| 95-99% | 666 | 336 | 2.39 (2.06 - 2.78) | 9.88×10<sup>-23</sup> | 73 | 1.92 (1.46 - 2.52) | 3.72×10<sup>-6</sup> | 137 | 2.76 (2.22 - 3.43) | 2.20×10<sup>-16</sup> | 87 | 2.17 (1.67 - 2.80) | 3.05×10<sup>-9</sup> | 0.103 |
| >99% | 149 | 102 | 3.25 (2.50 - 4.22) | 1.52×10<sup>-17</sup> | 27 | 3.17 (2.06 - 4.87) | 8.15×10<sup>-8</sup> | 32 | 2.88 (1.93 - 4.29) | 1.30×10<sup>-7</sup> | 29 | 3.23 (2.12 - 4.90) | 1.78×10<sup>-8</sup> | 0.916 |
| P Trend | 1.75×10<sup>-203</sup> |  |  |  | 2.34×10<sup>-45</sup> |  |  | 4.92×10<sup>-65</sup> |  |  | 2.57×10<sup>-41</sup> |  |  |  |

OR, Odds Ratio; Confidence Intervals, CI;

### Combined effect of PRS and family history

We then assessed the incremental contribution of adding wPRS_116_ and family history to a baseline model that included age, sex and the first 10 genetic PCs as predictors. We found no statistical interaction between the wPRS_116_ and sex, age, or family history (Table 5, P_interaction_ = 0·426 for multiplicative interaction with sex, P_interaction_ = 0·688 with age, P_interaction_ = 0·388 with family history), therefore we did not fit additional interaction terms in the model. A logistic regression on age, sex and the 10 PCs yielded a c-statistic of 0·527, and the corresponding estimate of Λwas 0·01 bits (Table 6). When adding family history alone, the c-statistic increased to 0·552 and the corresponding Λwas 0·02 bits. Adding both family history and wPRS_116_ yielded c-statistic of 0·610 and an incremental value of 0·10 bits, which showed significantly improvement over family history alone. When recalibrating the posterior probabilities, it showed no increases in the test log-likelihood and indicated the model (baseline + family history + wPRS_116_) was well-calibrated (Table 6).

**Table 5.** Interactions between PRS with age, sex and family history (FH).

|  | UK Biobank (n = 25 087) |  |  |
| --- | --- | --- | --- |
|  | beta | se | p |
| PRS | 0.056 | 0.003 | <2×10<sup>-16</sup> |
| PRS*sex | 0.004 | 0.005 | 0.426 |
| PRS*age | 0.0001 | 0.0004 | 0.689 |
| PRS*FH | -0.004 | 0.006 | 0.471 |

**Table 6.** Incremental contribution of genetic factors to CRC detection compared with family history (UKBB).

| Models | Crude<br>C-statistics | Model-based<br>C-statistics | Crude $\Lambda$<br>(bits) | Model-based<br>$\Lambda$ (bits) | Incremental | Difference in<br>Test log-likelihood (nats) | Difference in log-likelihood after<br>Recalibration (nats) |
| --- | --- | --- | --- | --- | --- | --- | --- |
| Baseline only | 0.527 | 0.527 | 0.01 | 0.01 | -- | -- | 2 |
| Baseline + FH | 0.552 | 0.553 | 0.03 | 0.03 | 0.02 | 70 | 1 |
| Baseline + wPRS<sub>116</sub> | 0.610 | 0.611 | 0.11 | 0.11 | 0.10 | 281 | 0 |
| Baseline + wPRS<sub>116</sub> +<br>FH | 0.610 | 0.611 | 0.11 | 0.11 | 0.10 | 281 | 0 |
| Baseline + regional<br>scores | 0.597 | 0.598 | 0.09 | 0.09 | 0.08 | 108 | 2 |

### Absolute risk of developing CRC by levels of PRS with age and sex

To gauge the potential public health impact of applying such risk prediction model in the general population, we estimated the 10-year absolute risk of the general UK population (Figure 2, appendix p 8 Table S5). We observed that the estimated absolute CRC risk for individuals at the highest 1% of PRS began exponentially to increase after 45 years old, and reached a threshold of 22·1% risk in men and 14·4% risk in women by 75 years of age. As 50 years of age is the recommended starting age of screening in the UK, we used the average risk at this age as the reference threshold (0·48% for man and 0·33% women). Individuals in the top 10% of wPRS_116_ would reach or exceed this level of risk at 45 years old, which is 5 years earlier than the average risk population; in contrast, individuals in the bottom 10% of PRS would stay below this average risk until 60 years old. The age lag to reach the same level of risk could be as much as 15 years. If we considered individuals with 10-year absolute risk ≥5% as high risk group, with risk strata by wPRS_116_ in population settings, we will able to identify 10% men and 5% women meriting intensive screening at 65 years old.

**Figure 1.**
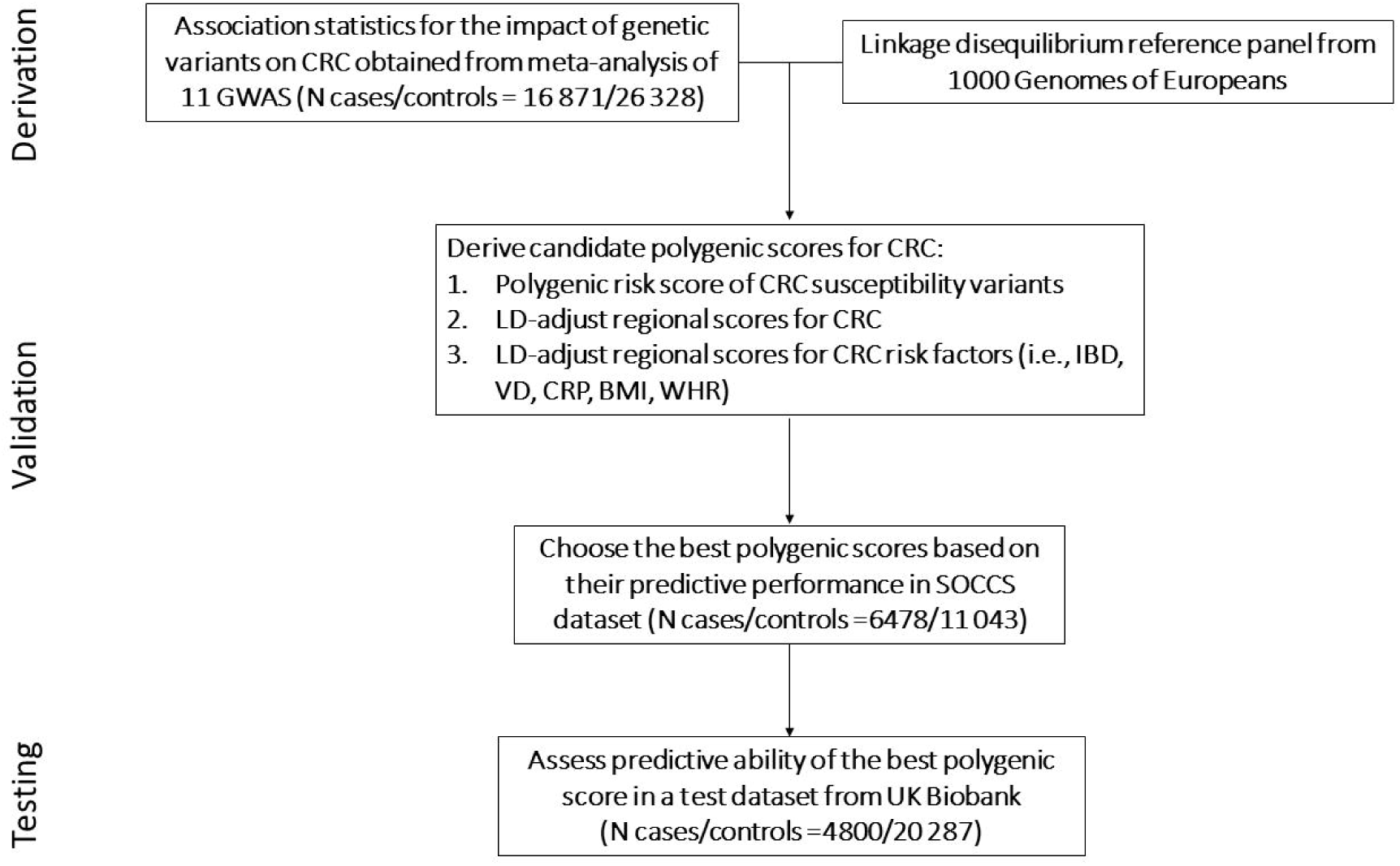
Schematic representation of the study design.

**Figure 2.**
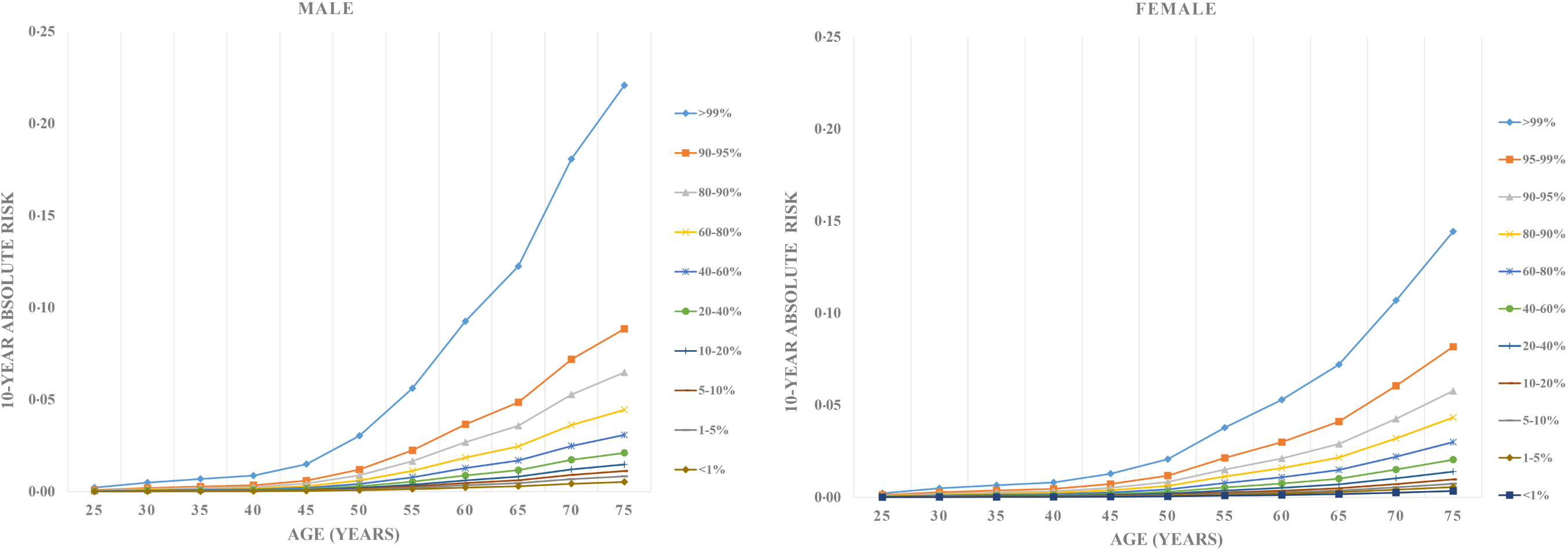
10-year absolute risk of developing CRC in men and women. Absolute risks were calculated based on the UK incidence and mortality rates and using the wPRS_116_ relative risk estimated as described.

## DISCUSSION

In this study we describe a systematic approach to derive, validate and test a number of candidate genetic risk scores with incorporating information from hundreds to thousands of common genetic variants to predict polygenic susceptibility of CRC. We evaluated the predictive performance of both a genomic risk score and a series of regional genetic scores that were built based on the summary statistics from multiple GWASs. Our study shows that a weighted genomic risk score including 116 CRC susceptibility SNPs is the optimal score with the strongest performance, while deconstructing genetic risk into multiple regional scores or inclusion of additional SNPs above the genome-wide significance threshold showed no further improvement on prediction performance. By implementing the optimal PRSs, we show that the inclusion of genetic factors into a baseline model of age, sex and family history results in a significant improvement of predictive power and opens up the potential for targeted screening for CRC risk.

The first objective of this study was to capture all CRC susceptibility SNPs and estimate the weights of the corresponding SNPs by meta-analyzing 11 studies with more than 42 710 individuals. The majority of the 116 SNPs had previously been identified but were further validated in this meta-GWAS without inclusion of study samples used for model development and evaluation. Although each variant individually exerts only modest effects on CRC risk (106 of 116 have per-allele odds ratio <1·20), the joint effect of SNPs as wPRS strengthens the association with an observed OR of 1·46 in Scottish dataset and 1·49 in UK Biobank dataset for per 1 SD increase of wPRS_116_.

ROC analysis of the genetic model that included wPRS_116_ showed an improved but still modest discriminative performance (c-statistic: 0·603 in Scottish dataset, 0·610 in UK Biobank dataset). To our knowledge, the best predictive performance achieved by PRS along with age and family history was 0·692 and 0·603 for Korean men and women,^22^ but the SNPs were chosen from the same dataset used to generate the model, and therefore the reported c-statistics are likely inflated. Other genetic models showed consistently low to modest discriminatory abilities. We previously developed a model including family history and 10 common genetic variants yielding a c-statistic of 0·56.^5^ Hsu *et al* developed sex-specific models by using family history and 27 common genetic variants with adjustment of endoscopy history and obtained a discrimination ability of 0·59 for men and 0·56 for women.^23^ Similarly, Smith *et al* reported a c-statistic of 0·57 for genetic risk model combing 41 CRC susceptibility SNPs.^24^ The most recent genetic model for CRC was developed by Jeon *et al* including 63 CRC susceptibility SNPs and achieved a very slightly improved predictive accuracy with a c-statistic of 0·59.^25^ This modest level of test performance is consistent across study populations, suggesting that risk assessment algorithms based on independent SNPs reaching genome-wide significance level have similar performance characteristics in European populations.

With the expectation of improving the predictive power of common genetic variants, we additionally derived a set of SNPs associated with CRC risk with liberal p-value thresholds to allow the contribution of signals from additional susceptibility SNPs that have not been discovered or validated in previous GWAS efforts. Any correlations between SNPs were addressed by creating LD-adjusted regional scores. However, with inclusion of thousands of SNPs, the predictive capacity did not improve but showed a lower c-statistic in the range of 0·58 to 0·59, which is probably due to the cost of adding noise from SNPs that were not truly associated with CRC. To assess if the genetic susceptibility of known risk factors of CRC would further contribute to CRC prediction, we developed prediction models, which incorporated genetic information of several known risk factors, but the c-statistic remained close to 0·60.

Most previous efforts mainly focused on the predictive ability of PRS to capture the overall risk of CRC.^5,6,23-25^ However, there is compelling evidence suggesting that genetic risk factors may differ by anatomic locations.^26,27^ We therefore specifically aimed to improve prediction of site-specific CRC by deconstructing the commonly used genomic risk score into several regional scores, allowing susceptibility signals through multiple/different mechanisms to influence genetic predisposition to site-specific CRC. Using proximal, distal and rectal cancer as distinct disease endpoints, we derived a subset of regional scores influencing cancer at each anatomic site, but their predictive performance still showed modest discriminative ability. Better annotation and characterization of these clusters of genomic regions on a biologically mechanistic basis would be helpful to capture genetic heterogeneity in site-specific CRC, but it is beyond the scope of the present study.

An extrapolation to the UK population led to the conclusion that 10% of the general population will have a 10-years absolute risk approaching 5% after 65 years old on the basis of quantifiable genetic risk alone and who will merit intensive screening. A 5% threshold of absolute risk has clinical and public health impact since it exceeds the highest risk at any age in the general population and it is 10-fold greater than the risk of a 50-year old person who is eligible to enter the population-based screening programs. Additionally, the modelling shows individuals at different levels of the wPRS_116_ will reach the same risk estimate at different ages, supporting the notion that using genetic profiling in combination with age will lead to more effective routine screening.

Estimates of absolute risk derived from our analysis may help to refine recommendations regarding the age at screening initiation. Although CRC family history allows identification of a small proportion of high risk individuals, there is considerable potential for recall bias and it does not allow any differentiation of risk among the vast majority of people without family history. Profiling CRC susceptibility by genetic testing would overcome such shortcomings and provides additional inheritable information that is not captured by family history. In addition, given that the screening participation rate in Scotland is ∼60%,^28^ it is possible that those identified to be at higher CRC risk by their genetic profile might be more likely to participate in screening, thereby further increasing the screening uptake and detection rates.^29,30^ Currently in Scotland the bowel screening programme uses the quantitative fecal immunochemical test (qFIT).^28^ Implementation of qFIT-based screening in conjunction with PRS-based risk profiling would achieve a fine-tuned sensitivity/specificity of the qFIT test. The application of a high cut-off qFIT threshold in those at low genetic risk could avoid invasive tests and minimize adverse effects and cost. Conversely, for high genetic risk groups, the sensitivity of qFIT can be increased by adjusting the cut-off concentration to a lower value.

By exploiting a wealth of data available to us, we conducted a rigorous 3-stage study with score derivation, optimization and evaluation in multiple datasets. We reduced the dimensionality of the prediction task from millions of common variants to a hundred CRC susceptibility SNPs and multiple regional genetic scores to develop models for both overall and site-specific CRC. It should be noted that the optimal GRS derived in this study was mainly based on the tagging variants in meta-GWAS, while application of underlying functional or causal variants with potentially stronger effect may further improve the risk discrimination. For site-specific models, although we treated proximal, distal and rectal cancer as distinct endpoints to generate the best set of regional scores respectively, the weights used for score calculation were derived from the coefficient estimates for overall CRC. Although we have increased the number of CRC susceptibility SNPs significantly (to >100 SNPs), the contribution to individualized risk profiling is limited. For the purposes of personal profiling, the optimal PRS needs to be combined with information on the external (e.g., lifestyle) and internal environment (e.g., microbiome) and with realistic measures of clinical status (e.g., biomarkers) to provide an integrated description of individual trajectories from different dimensions.

## Data Availability

All data referred to is available in the manuscript.

## Acknowledgement

We are grateful to all who contribute to recruitment, data collection and data curation. We acknowledge that these studies would not be possible without the patients and controls and their families. We acknowledge the expert support on sample preparation from the Genetics Core of the Edinburgh Wellcome Trust Clinical Research Facility.

## Contributors

ET and MD conceived the study; XL undertook data manipulations and statistical analysis with input from AS and PM; MT performed the meta-GWAS analysis. HC, RSH, IPMT, SMF, YH, XZ and MGD provided access to GWAS data. XL wrote the article with input from other authors. All authors critically reviewed the manuscript and contributed important intellectual content. All authors have read and approved the final manuscript as submitted.

## Ethics committee approval

Ethics approval and consent to participate SOCCS received ethical and management approvals from the Multi-Centre Research Ethics committee for Scotland (approval number MREC/ 01/0/5). UK Biobank has approval from the North West Multi-Centre Research Ethics Committee (11/NW/0382) and obtained written informed consent from all participants prior to the study.

